# Echocardiography-Based Deep Learning Model to Differentiate Constrictive Pericarditis and Restrictive Cardiomyopathy

**DOI:** 10.1101/2022.11.29.22282900

**Authors:** Chieh-Ju Chao, Jiwoong Jeong, Reza Arsanjani, Kihong Kim, Chadi Ayoub, Martha Grogan, Garvan Kane, Imon Banerjee, Jae K Oh

## Abstract

**Background:** Constrictive pericarditis (CP) is an uncommon but reversible cause of diastolic heart failure if appropriately identified and treated. Although echocardiography can detect CP based on characteristic cardiac motion and Doppler findings, its diagnosis remains a challenge for clinicians. Artificial intelligence (AI) may enhance identification of CP. We proposed a deep learning approach based on transthoracic echocardiography (TTE) to differentiate CP from restrictive cardiomyopathy (RCM).

**Methods:** Patients with a confirmed diagnosis of CP and cardiac amyloidosis (CA, as the representative disease of RCM) at Mayo Clinic Rochester from 1/2003-12/2021 were identified to extract baseline demographics and the apical 4 chamber (A4C) view from TTE studies. The cases were split into a 60:20:20 ratio for training, validation, and held-out test sets of the ResNet50 deep learning model. The model performance (differentiating CP and CA) was evaluated in the test set with the area under the curve (AUC). GradCAM was used for model interpretation.

**Results:** A total of 381 patients were identified, including 184 (48.3%) CP, and 197 (51.7%) CA cases. The mean age was 68.7±11.4, and 72.8% were male. ResNet50 had a performance with an AUC to differentiate the 2-class classification task (CP vs. CA, AUC 0.97). The GradCAM heatmap showed activation around the ventricular septal area.

**Conclusion:** With a standard A4C view, our AI model provides a platform for the early and accurate detection of CP, allowing for improved workflow efficiency and prompt referral for more advanced evaluation and intervention of CP.

## Introduction

Constrictive pericarditis (CP) is an uncommon but reversible cause of diastolic heart failure if properly identified and treated ^1–3^. In Western countries, the etiology of CP cases was reported as idiopathic or viral-induced, followed by cardiac surgery and prior chest radiation^4,5^. The clinical presentation of CP can be similar to that of a myocardial disease or could be misdiagnosed as gastrointestinal disorder or hepatic failure due to predominant right-hear failure symptoms such as hepatic congestion or ascites^2,6^. Hatle et al first described characteristic TTE and Doppler findings of CP and our group established the echocardiographic diagnostic criteria of CP ^7^, which largely facilitated the diagnosis of the disease. The criteria include characteristic ventricular septal motion, augmented septal mitral annulus motion, and restrictive mitral diastolic filling with respiratory variation. However, even with the echocardiographic diagnosis criteria^8^, correctly interpreting echocardiographic signs requires a high level of training and experience. Thus the diagnosis remains a challenging task for clinicians ^1,9^. Despite the implementation of multi-modality cardiac imaging as a diagnostic tool in the past decade, misdiagnosis or delayed diagnosis is not uncommon ^6,10–12^.

With the advance of artificial intelligence (AI) technology, its introduction to clinical workflow has been proposed to facilitate the diagnosis of CP^13,14^. In 2016, Sengupta et al. undertook a pilot study using speckle-tracking strain data to differentiate CP from restrictive cardiomyopathy (RCM), but this approach was based on an associate memory classifier instead of end-to-end deep learning^15^. Deep learning algorithms allow direct extraction of features from the raw images, may enable superior performance, and have been widely applied in echocardiography-based studies for disease classification tasks ^16–19^. While a deep learning model that identifies CP would be clinically beneficial, such a model has not been established yet. This is owing to the rarity of CP cases that can be used for model training and the high-level expertise in echocardiography required for correct diagnosis^15^.

Deep learning models could be overfitted when only a small sample size is available for training^20^. To address the training sample size limitation in diseases with a lower prevalence, our group proposed a frame-based approach for echocardiography data augmentation^21^. In this work, we proposed a relatively simple deep learning approach based on the standard apical 4 chambers (A4C) TTE view to differentiate CP from RCM since A4C view shows ventricular septal motion, mitral annulus motion, and left ventricular filling pattern.

We hypothesized that the deep learning model could accurately differentiate CP from RCM based on the standard A4C view from TTE studies. Cardiac amyloidosis (CA), a type of RCM due to the infiltrative process of amyloid proteins in the myocardium was chosen as the representative disease of RCM^25–27^. We anticipate the proposed model can facilitate the diagnosis of CP, and further promote the application of echocardiography-based deep learning models in rare diseases.

## Method

### Patient population

The study was approved by the Mayo Clinic Institutional Review Board (protocol #19-009303). Patients who underwent a TTE study at Mayo Clinic Rochester from 01/01/2003 through 12/31/2021 were reviewed to identify the cases of CP and CA (as the representative of RCM). Specifically, the diagnosis of CP was confirmed by surgery, and the diagnosis of CA was established by endomyocardial biopsy or advanced imaging modalities^26^. Patients were excluded if any of the following conditions were present in the echocardiographic study:

1. 2D study: Patients with inadequate echocardiographic images, ≥ moderate aortic/mitral regurgitation or aortic/mitral stenosis, significant pericardial effusion, presence of prosthetic valve, mitral/tricuspid valve annuloplasty, conduction delay (≥ 1st degree AV block, left bundle branch block, AV dissociation), intracardiac device such as pacemaker, cardiac resynchronization therapy device or implanted cardioverter device. Only non-contrast images were selected.
2. Doppler study: Patients with increased respiratory effort (i.e. chronic obstructive lung disease, severe obesity), significant pericardial effusion, atrial fibrillation/flutter, severe RV dysfunction. If any of the 3 parameters (hepatic vein, mitral inflow, medial e’) was not available, patients were excluded.

The final cohort was manually labeled as CP and CA accordingly.

#### Image Pre-processing and Data Augmentation

The preprocessing block is similar to the preprocessing procedure described in our previous paper where the raw DICOM file of the A4C TTE video clips is loaded, individual frames are separated, and saved as .png files for further processing^21^. Once all the frames of various aspect ratios and image sizes were extracted, the frames were then converted to grayscale images and center cropped by removing the top and bottom 10% and the left and right 25% of the image.

After the cropping step, the images were matched to the average histogram of the training dataset and resized to the input of the model. The resized images then underwent additional steps in the pre-processing block, including respiratory line augmentation, consecutive frame stacking for wall motion embedding, color jitter, and gaussian noise augmentation. By exporting the individual frames, the sample size was increased with a factor of ∼ 95 (from 381 to 36,291).

#### Assimilating heart motion

Concerning a frame-based approach that may not be able to capture the temporal-spatial relationships in the characteristic septal shift of CP, we also attempted to incorporate the wall motion information for the model training. To embed motion information into the model training, we combined three consecutive frames into an RGB image where the non-grayscale color represents the motion information, as seen in **Figure 1**. The model performance was tested in images with and without RGB motion embedding.

**Figure 1.**
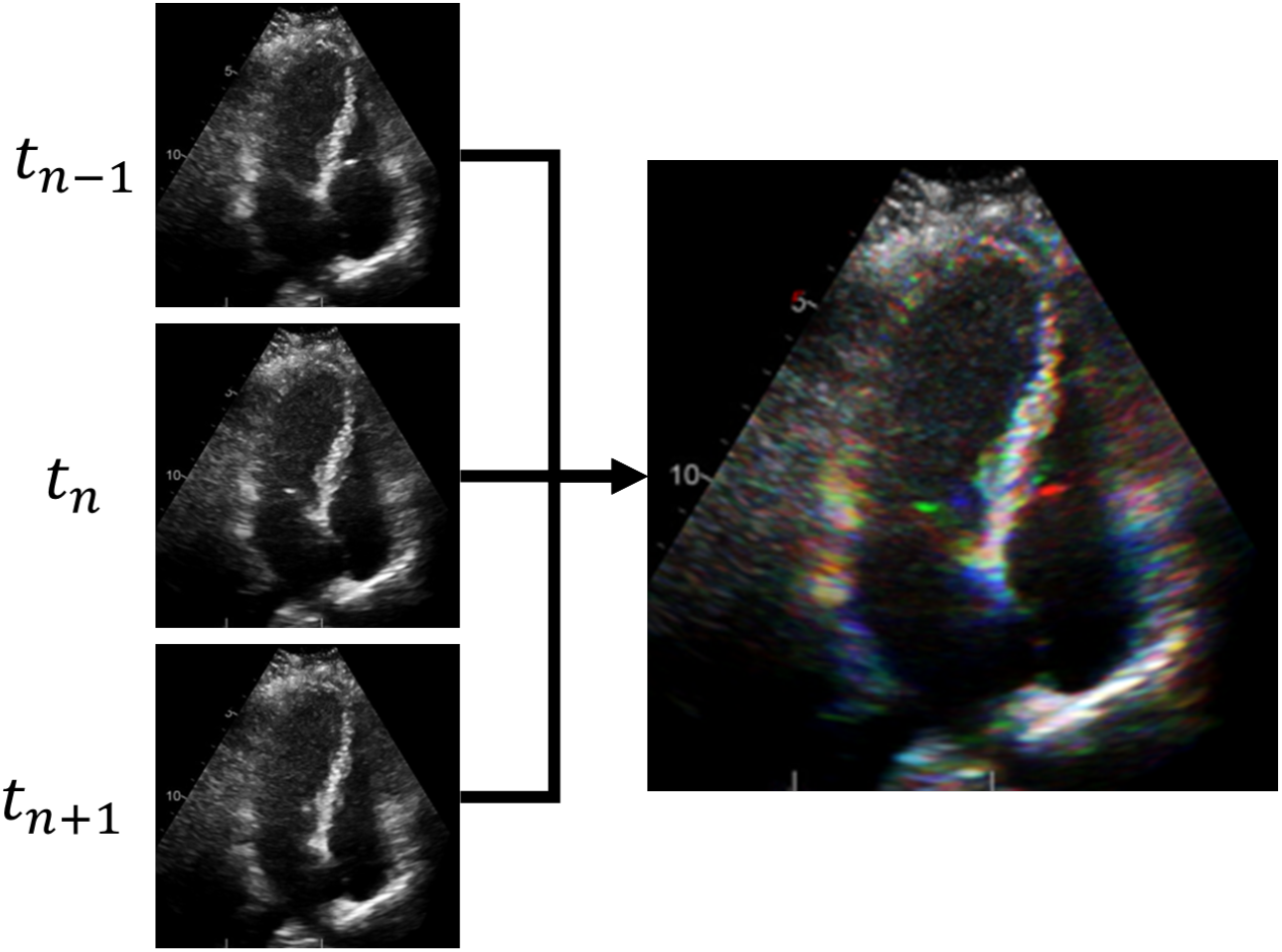
RGB motion embedding. For a given grayscale frame in the study, the previous frame (t _n-1_) and the next frame (t _n+1_) were used to create an RGB image that contains 3 channels of data. This RGB motion embedding procedure is anticipated to contain the wall motion information within a still image instead of a full video clip.

#### Framework

**Figure 2.** is a diagrammatic representation of the proposed framework that highlights the core processing blocks, namely the preprocessing block, framewise classification block, and study-level diagnosis block (meta-learner). In brief, the trained framework is designed to directly read the video clips and produce a probabilistic diagnosis at the exam level. The classification block tested ResNet50 that were deemed to be relevant to the classification task. Finally, a meta-learner block consolidates the frame-level predictions into one study-level prediction.

**Figure 2.**
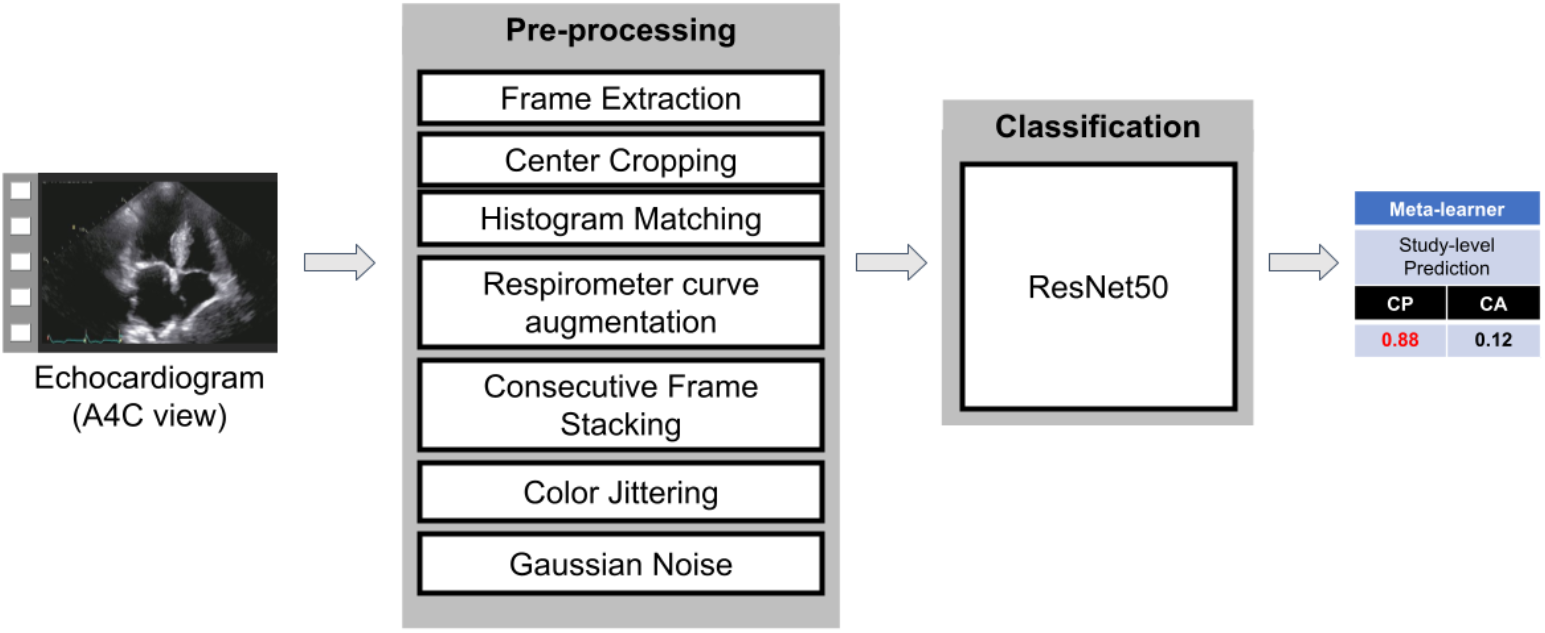
The architecture of the proposed model. Blocks represent the different modules.

#### Model Selection and Training

The ResNet50 was selected as the candidate model ^28^. The hyperparameters used for the consecutive frame classification method were a batch size of 32, a learning rate of 0.000001, and a weight decay of 0.3 with cross-entropy loss for 200 epochs. The models were trained on an Nvidia RTX A5000 GPU. To prevent data leakage, we generated the split at the study level so that no images from the same study are mixed between the train and the test sets. The models were trained for a binary (CP and CA) classification task. We evaluated the quantitative performance of each model in terms of area under the receiver-operating characteristic curve (AUC ROC), precision, recall, and F1-score.

#### Model interpretability: GradCAM and ablation studies

Ablation procedures were performed to test the 2-class model’s performance with different input data, as described in **Figure 3**. GradCAMs are often used to interpret the model’s performance by localizing the activations of the final convolution layer of the model during inference^29^. Following these revelations, we combined GradCAM with ablation studies, that ablate parts of the image, to understand where in the image the model is basing its predictions.

**Figure 3.**
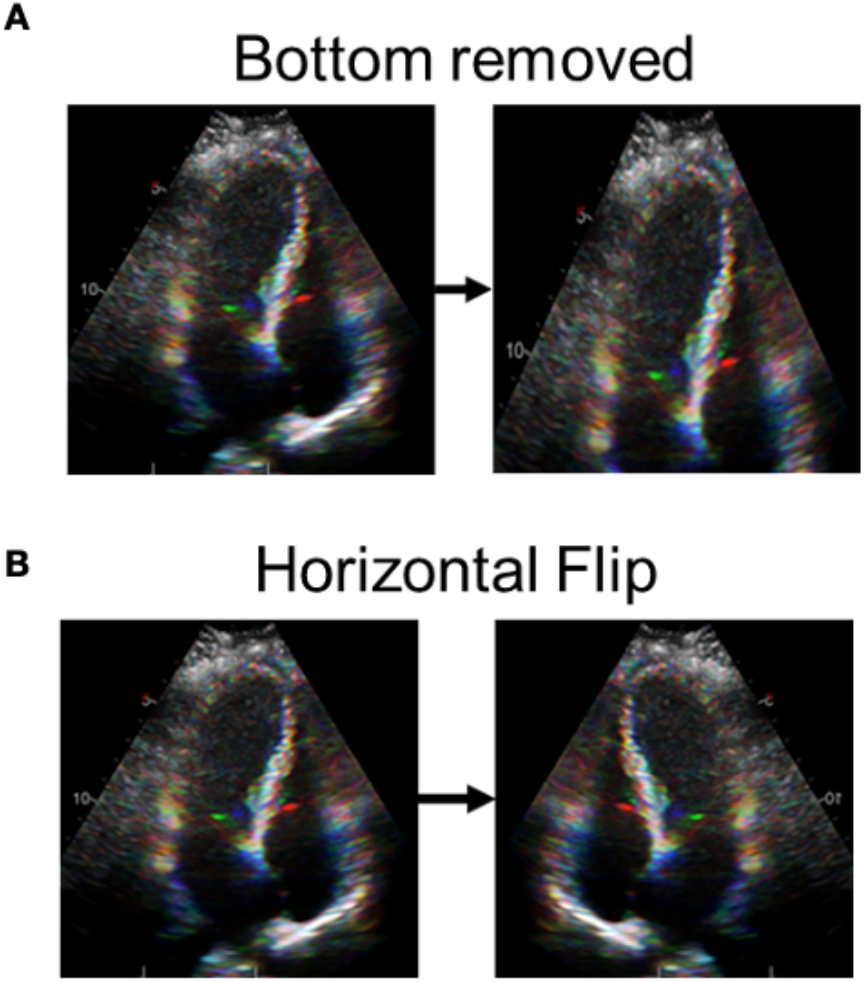
Ablation procedures performed in this study. **Panel A**. The lower one-third portion of the A4C view may contain an ECG signal and the respirometer line so was removed to test whether the model was depending on this information. **Panel B**. Horizontal flip was conducted to simulate the A4C view format at different institutions.

**Figure 3.**
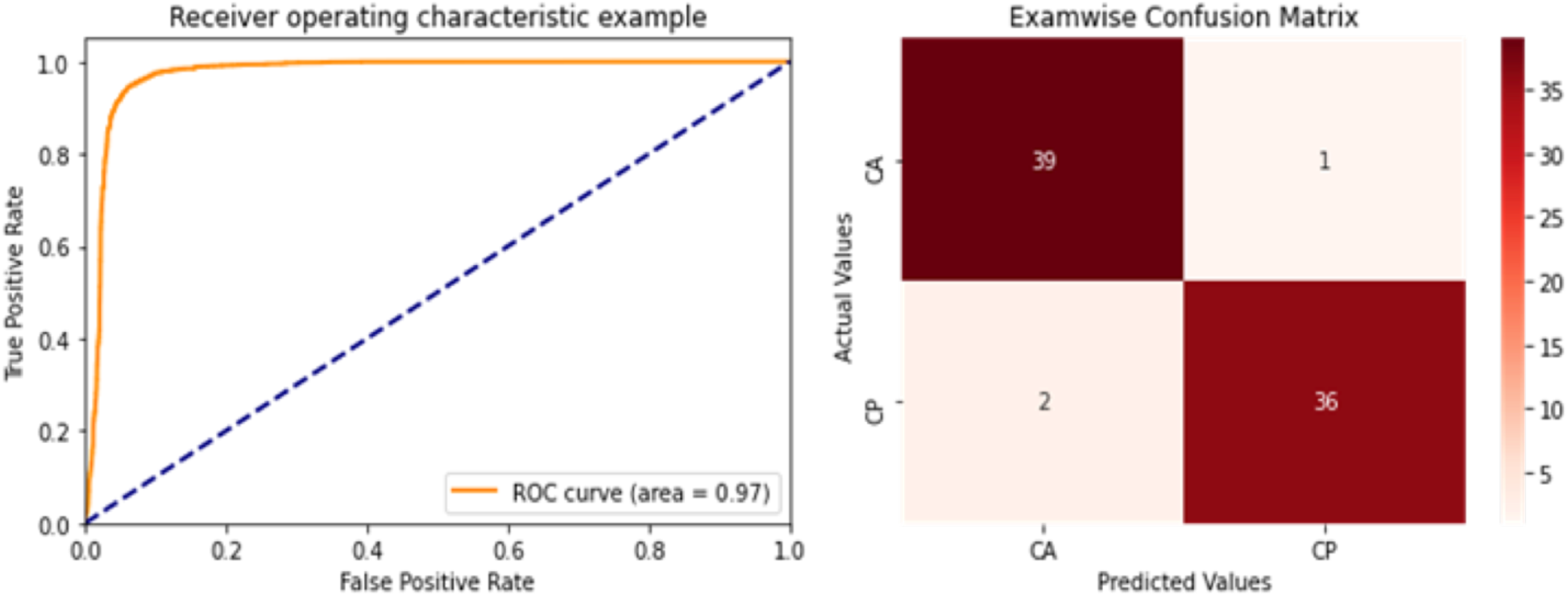
ResNet50 Model Performance for 2-class Classification task (CA and CP). Model performance was summarized as AUC of ROC (left) and confusion matrix (right) in each panel. The AUC ROC was 0.97.

## Results

### Patient Baseline Demographics

The final cohort contains 381 patients, of which 184 patients were CP, and 197 patience were CA. The mean age was 68.7± 11.4 years, and 72.8% were male. Detailed baseline patient characteristics are summarized in **Table 1**.

**Table 1.** Baseline Patient Characteristics.

| Characteristics | Internal Train and test |
| --- | --- |
| *Total study* | 381 |
| *No. of frames* | Total: 36,291<br>Train: 21,186<br>Validation: 7,717<br>Test: 7,388 |
| *Age (mean±SD)* | 68.7±11.4 |
| *Gender* | Male: 72.8%<br>Female: 27.2% |
| *Race /* | White: 86%<br>Asian: 10.5%<br>Black: 3.4% |
| *Ethnicity* | Hispanic or Latino: 14%<br>Not Hispanic or Latino: 86% |
| *Comorbidities at the time of TTE* | AFib: 59 (15%)<br>Cancer: 6 (2%)<br>Hypertension: 109 (29%)<br>Coronary Artery Disease: 59 (15%)<br>Chronic Kidney Disease: 75 (20%)<br>Diabetics (Type I and Type II): 37 (10%) |
| *Diagnosis*<br>*Cardiac Amyloidosis (CA)*<br>*Constrictive pericarditis (CP)* | CA: 197 (51.7%)<br>CP: 184 (48.3%) |

#### Comparison of single frame vs motion-embedded RGB image

While the model was able to extract information about the pericardium from the images from single frame images and perform well on the test set, our initial experiments showed that adding the motion information significantly improved the performance of the model’s recall across all classes and on average, improved the performance across all classes.

#### Quantitative performance

The model performance (precision, recall, and F1-score) was summarized in **Table 3**. The study-level AUCROC and confusion matrix were shown in **Figure 3**.

#### GradCAMs and ablation study results

**Figure 4.** showed the representative GradCAMs of the 2-class models. Overall, the activation of GradCAM was focused on the septal regions for prediction. While the activation area slightly moved in the ablation studies (**Figure 4, Panel C** and **D**), the model performance was similar. Details are summarized in **Table 5**.

**Figure 4.**
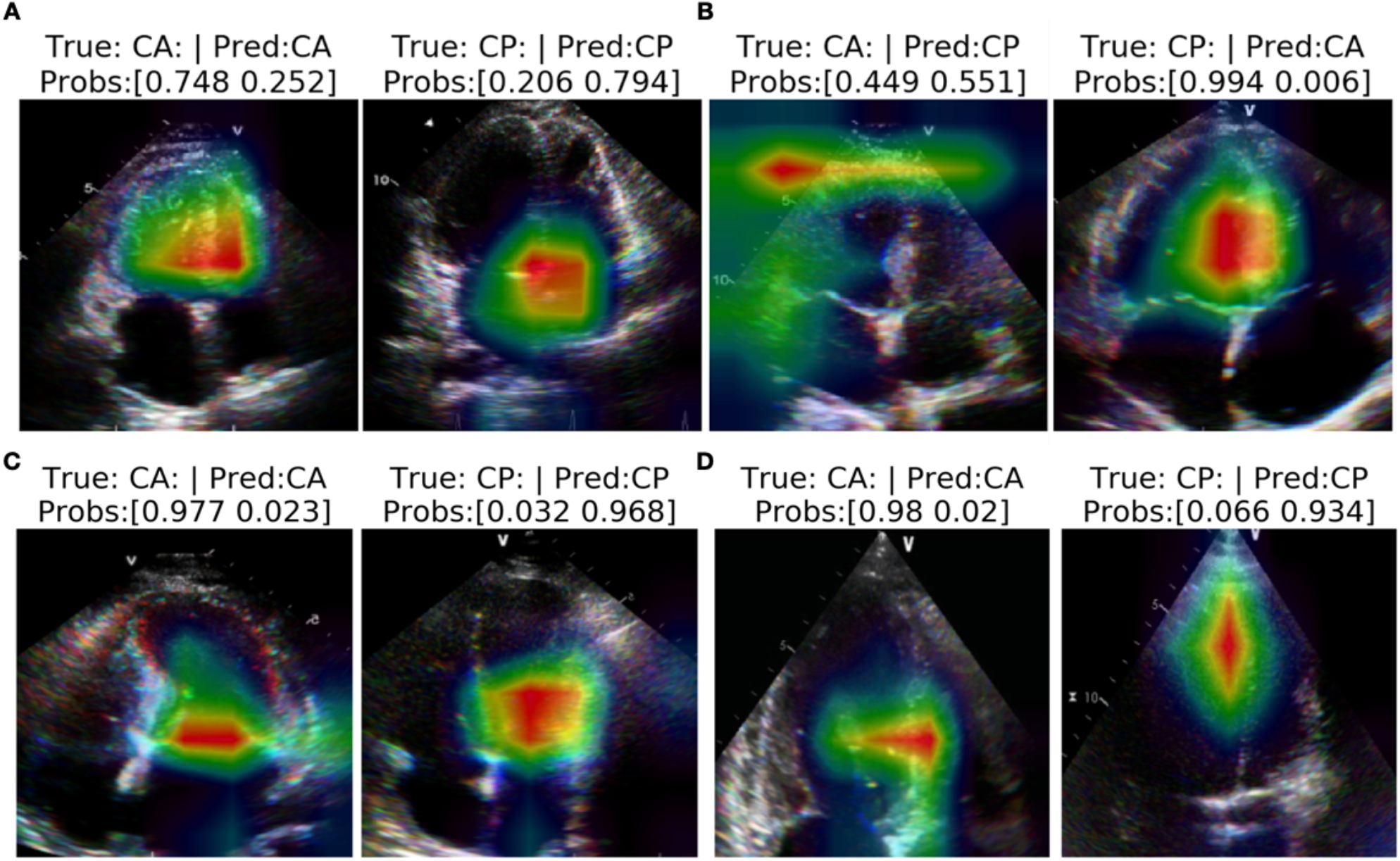
Representative GradCAMs. The GradCAM activation was mainly around the basal-to-mid septal area in correctly classified (**Panel A**) and incorrectly classified (**Panel B**) cases. **Panel C**. demonstrated the GradCAM in horizontally flipped images, which still covered the septal area but had some focus on the mitral valve area. **Panel D**. With the bottom third removed, the activation area slightly moved distally to the mid-LV region.

## Discussion

In this retrospective study, we successfully developed the first echocardiography-based deep learning model which can accurately differentiate CP from CA (as a representative of RCM). The major strengths of this work include 1) the excellent overall model performance (ResNet50, AUC 0.97) in differentiating CP and CA using only the standard A4C view and 2) the frame-based approach with embedded wall motion information provided a computationally efficient option which avoided overfitting and preserved the spatial-temporal relationship in the meantime. We foresee the potential of this model to enable an automated clinical workflow to improve the quality of interpretation and facilitate the diagnosis of CP, especially at institutions without trained echocardiography experts. Furthermore, we hope our approach can facilitate future echo-based studies for other uncommon conditions like CP.

### Facilitating the Diagnosis of CP with Artificial Intelligence

Echocardiography has been considered the first-line diagnostic tool for CP^6,8^. However, the accuracy of echo-based diagnosis largely depends on the quality of the echocardiography study, which requires skilled sonographers and experienced interpretation physicians^6^. Even at a tertiary center, correctly establishing the diagnosis of CP can be challenging in complex cases, and may require multi-modality imaging or invasive hemodynamic studies ^24,30,31^.

A majority of published CP studies were from tertiary referral centers due to the rarity and complexity of CP^2,3,7,8,32^. Prior to our study, Sengupta et al. was the only group that reported the effectiveness of a machine learning approach for CP. Their work incorporated 15 variables, including heart rate, speckle tracking strain data from A4C, and short axis views, and reached an average AUC of 0.892 ^15^. Adopting the concept of automation and acceleration with a machine learning approach^15^, we further explored the use of directly extracting features with deep learning algorithms with a fully automated process and achieved an overall AUC of > 0.95.

Additionally, our work suggested that the A4C view contains most of the necessary information for the models to make correct predictions. This was implied in Sengupta’s work that 13 out of their 15 final input variables were obtained from the A4C view^15^.

If incorporated into clinical workflow, we foresee this model simplifying the echocardiography laboratory workup for CP, and further improving the quality of interpretation and facilitating diagnosis. This model can be especially beneficial in institutions without trained echocardiography experts^15^. In current practice, to obtain the required images and Doppler parameters for diagnosis, echocardiography workup for CP requires skilled sonographers to scan over the respiratory cycle^8^. Furthermore, interpreting physicians need to review multiple long clips before making the diagnosis of CP. In contrast to the above time-consuming process, provided the standard A4C view is available, our model can be combined with real-time system-generated alert messages to suggest the presence of CP and avoid misdiagnosis. Our model provides an option to improve the efficiency of echocardiography laboratories’ workflow and potentially prompt early referrals to tertiary centers for more advanced evaluation and intervention of CP.

### AI-driven insight for the diagnosis of CP

In the proposed echocardiographic diagnostic criteria for CP by Welch et al., the principal variables that are independently associated with CP were mainly obtained from the A4C view^8^. Specifically, the most important feature is the presence of ventricular septal shift, which can be combined with either medial e’ or hepatic vein diastolic reversal signal to reach an optimal sensitivity and specificity profile. Importantly, using different combinations of the criteria can vary the sensitivity and specificity^8^. This reflected the limitations of conventional approaches, which often rely on binary variables without incorporating conditional probabilities. In the current work, we omitted the signal from the hepatic vein but still reached an overall superior performance. This finding was consistent with the recent machine learning study using strain data^15^.

The GradCAM analysis (**Figure 4**) showed that our model was overall focusing on the septal area, suggesting that the model was analyzing features related to septal bounce. In the ablation study, we removed the lower one-third images (to remove the respirometer curves) but still observed a similar model performance and heatmap demonstrated by GradCAM (**Figure 4D**). These results also confirmed that the model was not relying on the respirometer curves to recognize CP.

Additionally, while the spatial-temporal relationship was not incorporated in a standard framed-based approach (in contrast to video clips), this issue was addressed by embedding heart motion into RGB images. Using the motion embedding RGB images successfully captured the septal bouncing feature and improved model performance, especially the recall (sensitivity) (**Table 2**). Our frame-based approach can be especially useful when developing models for rare diseases, which avoids overfitting with limited training video samples but still allows the preservation of spatial-temporal relationships.

**Table 2.** Comparison between single-frame and multi-frame (RGB motion embedding) trained models.

|  | Single-Frame Test |  |  | Multi-Frame Test |  |  |
| --- | --- | --- | --- | --- | --- | --- |
|  | Precision | Recall | F1-score | Precision | Recall | F1-score |
| **CA** | 0.911±0.045 | 0.900±0.005 | 0.905±0.023 | 0.924±0.038 | **0.925±0.004** | 0.924±0.019 |
| **CP** | 0.913±0.044 | 0.938±0.003 | 0.925±0.023 | 0.936±0.032 | **0.948±0.003** | 0.942±0.017 |

\*Performance metrics reported are bootstrap confidence intervals with 1000 iterations and a minimum sample size of 25% of the data. \*\*Boded results are significantly different than their counterparts with at least one standard deviation difference.

**Table 3.** Frame- and Study- Level ResNet50 Model performance.

|  | Frame-level Test |  |  | Study-level Test |  |  |
| --- | --- | --- | --- | --- | --- | --- |
| Class | Precision | Recall | F1-score | Precision | Recall | F1-score |
| CA | 0.924±0.038 | 0.925±0.004 | 0.924±0.019 | 0.946±0.042 | 0.976±0.024 | 0.961±0.025 |
| CP | 0.936±0.032 | 0.948±0.003 | 0.942±0.017 | 0.971±0.030 | 0.949±0.035 | 0.959±0.023 |

**Table 5.** Frame- and study- level model performance for 2-class classification task.

|  | Frame-level |  |  | Study-level |  |  |
| --- | --- | --- | --- | --- | --- | --- |
|  | Ablate Bottom Third of Frame (same model) |  |  |  |  |  |
| Class | Precision | Recall | F1-score | Precision | Recall | F1-score |
| CA | 0.879±0.058 | 0.892±0.005 | 0.885±0.031 | 0.941±0.045 | 0.899±0.045 | 0.918±0.034 |
| CP | 0.908±0.045 | 0.916±0.004 | 0.912±0.023 | 0.886±0.069 | 0.948±0.035 | 0.914±0.042 |
|  | Horizontal Flip |  |  |  |  |  |
| Class | Precision | Recall | F1-score | Precision | Recall | F1-score |
| CA | 0.870±0.060 | 0.902±0.005 | 0.884±0.032 | 0.915±0.056 | 0.899±0.044 | 0.906±0.037 |
| CP | 0.915±0.043 | 0.907±0.004 | 0.910±0.022 | 0.885±0.066 | 0.920±0.041 | 0.900±0.040 |

## Limitations

The study is limited by its retrospective nature. It is also subjected to referral bias as a tertiary center cohort. External validation data were not available, which is partly due to the rarity of CP cases as indicated above. While the overall training sample size is relatively small from a machine learning perspective, our cohort has contained one of the largest CP series available.

The frame-based approach with RGB motion embedding countered the issue of overfitting while preserving the spatial-temporal relationship. Potential target leak from the respirometer curve, and the limitation of generalization (Mayo format A4C view) were also addressed in our data augmentation process. Importantly, this model was specifically designed to differentiate CP from CA, the efficacy of this approach in more general cases was not investigated in this study.

## Conclusion

This work reported the first deep learning model with excellent performance for diagnosing CP. With a standard A4C view, our model provides an option to improve the efficiency of echocardiography laboratories’ workflow and potentially prompt early referrals to tertiary centers for more advanced evaluation and intervention of CP.

### Clinical Perspectives

#### Core Clinical Competencies

The diagnosis of CP has been a challenge for clinicians. While various approaches have been proposed, only one machine learning study is available for the diagnosis of constrictive pericarditis. We report the first deep learning model with excellent performance for diagnosing CP.

#### Translational Outlook

With a standard A4C view, our model provides an option to improve the efficiency of echocardiography laboratories’ workflow and potentially prompt early referrals to tertiary centers for more advanced evaluation and intervention of CP.

**Central Illustration.**
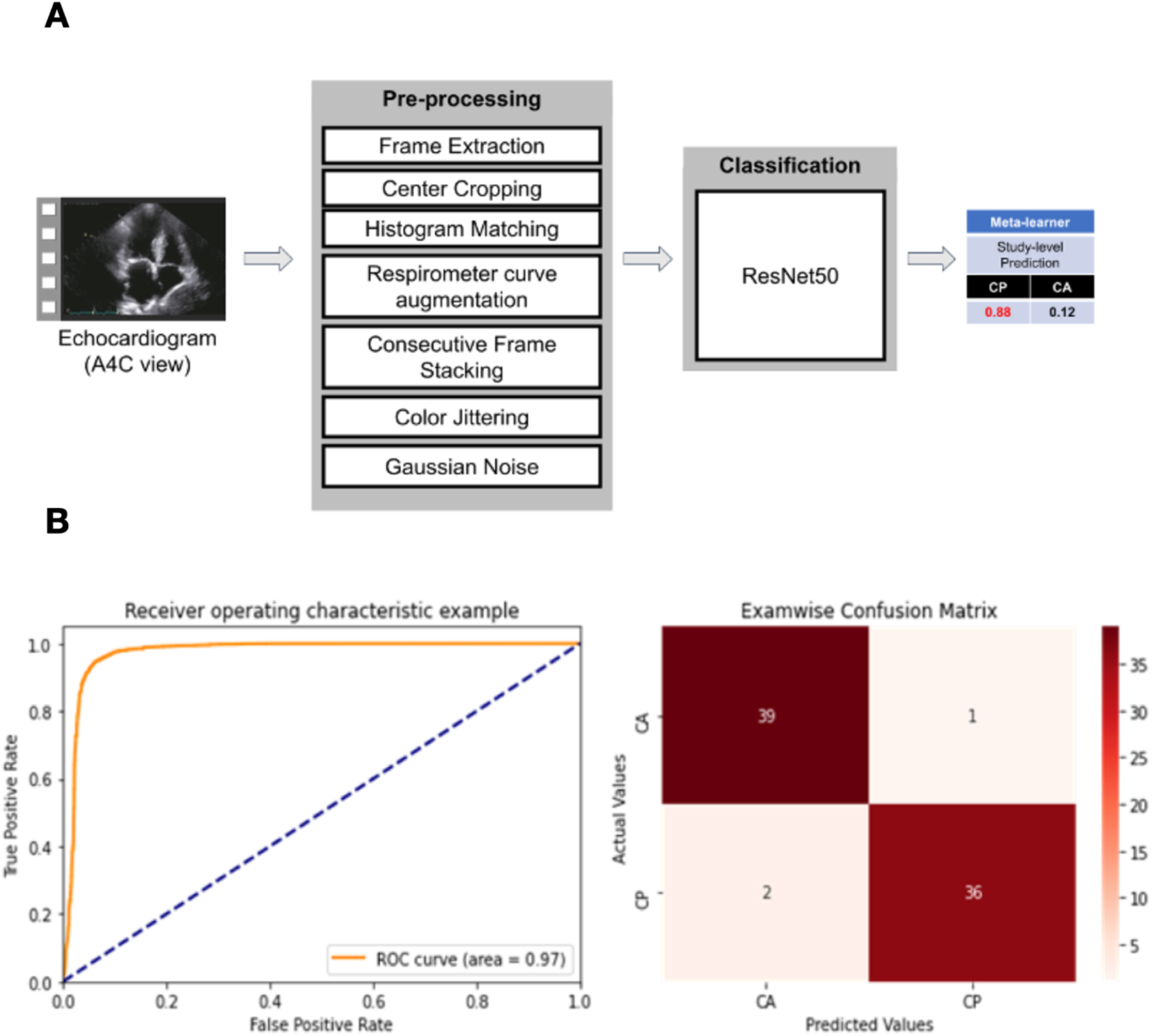
**Panel A.** Standard apical 4 chamber view (A4C) was used to train a ResNet50 model for differentiating constrictive pericarditis (CP) and cardiac amyloidosis (CA, as the representative of restrictive cardiomyopathy). **Panel B**. The model has excellent performance to differentiate CP and CA cases (area under the curve 0.97).

## Data Availability

All data produced in the present work are contained in the manuscript

## Abbreviations

A4C: apical 4 chamber view
AI: Artificial Intelligence
AUC: area under the curve
CA: cardiac amyloidosis
CP: constrictive pericarditis
RCM: restrictive cardiomyopathy
TTE: transthoracic echocardiography

